# Automated Detection of Early-Stage Dementia Using Large Language Models: A Comparative Study on Narrative Speech

**DOI:** 10.1101/2025.06.06.25329081

**Authors:** Kevin Mekulu, Faisal Aqlan, Hui Yang

## Abstract

The growing global burden of dementia underscores the urgent need for scalable, objective screening tools. While traditional diagnostic methods rely on subjective assessments, advances in natural language processing offer promising alternatives. In this study, we compare two classes of language models—encoder-based pretrained language models (PLMs) and autoregressive large language models (LLMs) for detecting cognitive impairment from narrative speech. Using the DementiaBank Pitt Corpus and the widely used Cookie Theft picture description task, we evaluate BERT as a representative PLM alongside GPT-2, GPT-3.5 Turbo, GPT-4, and LLaMA-2 as LLMs. Although all models are pretrained, we distinguish PLMs and LLMs based on their architectural differences and training paradigms. Our findings reveal that BERT outperforms all other models, achieving 86% sensitivity and 95% specificity. LLaMA-2 follows closely, while GPT-4 and GPT-3.5 underperform in this structured classification task. Interestingly, LLMs demonstrate complementary strengths in capturing narrative richness and subtler linguistic features. These results suggest that hybrid modeling approaches may offer enhanced performance and interpretability. Our study highlights the potential of language models as digital biomarkers and lays the groundwork for scalable, AI-powered tools to support early dementia screening in clinical practice.

## I. Introduction

**N**Eurocognitive disorders, including Alzheimer’s, Parkinson’s, and Huntington’s diseases, are becoming increasingly prevalent global health challenges. These conditions lead to significant declines in cognitive functions such as language, perception, attention, and memory. Currently, approximately 55 million people worldwide are living with dementia, and this number is expected to rise to 139 million by 2050 [1].

Early diagnosis of dementia is crucial for effective treatment and improving patients’ quality of life. However, traditional diagnostic tools—such as the Mini-Mental State Examination (MMSE), Mini-Cog Test, and Montreal Cognitive Assessment (MoCA)—have notable limitations:

- **Subjectivity and Bias [2]:** These assessments can be influenced by factors like educational background, cultural differences, and examiner bias, potentially leading to misdiagnoses.
- **Time and Cost Efficiency [3], [4]:** Multiple consultations and evaluations make the diagnostic process time-consuming and expensive, placing a burden on both patients and healthcare systems.

Given these challenges, there is a pressing need for innovative, objective, and efficient diagnostic methods. Advances in Artificial Intelligence (AI), particularly in Natural Language Processing (NLP), offer promising alternatives [5]. AI models can analyze linguistic patterns in patients’ speech to detect subtle signs of cognitive decline, enabling earlier and more accurate diagnoses [6].

**Large Language Models (LLMs)** like GPT-3 and GPT-4 have revolutionized NLP by effectively capturing complex language patterns and contextual nuances. Similarly, **Pre-trained Language Models (PLMs)** like BERT excel in understanding language representations for tasks such as classification and recognition. These models can identify subtle differences in speech that indicate early cognitive impairments, making them suitable for dementia detection.

In this study, we explore the effectiveness of various AI models in detecting dementia through linguistic analysis. Our key contributions are:

1. **Systematic Comparison of Language Model Architectures for Dementia Detection:** While previous studies have typically focused on single model types, we present the first comprehensive comparison between PLMs and LLMs for dementia detection. By evaluating BERT against modern LLMs (GPT-2, GPT-3.5 Turbo, GPT-4, and LLaMA-2), we provide crucial insights into their relative strengths and limitations in clinical applications.
2. **Novel Application of LLMs such as GPT-2, GPT3.5 Turbo, LLaMA-2, GPT3.5 Turbo and GPT-4:** While previous studies have extensively examined models like BERT and GPT-3, our study is the first to evaluate GPT2, GPT3.5 Turbo, LLaMA-2 and GPT-4 for dementia detection. This adds to the existing body of knowledge by exploring newer architectures and their potential in clinical diagnostics.

The rest of the paper is organized as follows: Section II reviews related work and provides background on dementia detection using AI models. Section III describes our methodology, including data collection, preprocessing, and model finetuning. Section IV details the experimental design. Section V presents and discusses the results of our experiments, and Section VI concludes the paper.

## II. Research Background

Dementia is a neurocognitive disorder that impairs cognitive function, including memory, thinking, and problem-solving. It is a chronic and progressive condition that can lead to significant disability and death. Alzheimer’s disease (AD) is the most common form of dementia, accounting for 60-80% of all cases [1]. Early detection of dementia is crucial for timely intervention and improved patient outcomes. However, traditional diagnostic procedures, such as clinical interviews and paper-based neuropsychological tests, have their limitations. They can be time-consuming, subjective, and inaccurate, especially in the early stages of the disease.

In recent years, the application of Natural Language Processing (NLP) techniques, particularly the fine-tuning of large language models (LLMs) and pre-trained language models (PLMs), has emerged as a promising approach in the early detection and classification of Alzheimer’s disease and related dementias (ADRD). This review examines recent advancements in utilizing models such as BERT, GPT variants, and LLaMA for dementia classification, highlighting their methodologies, performance, and limitations. Prior work has explored interpretable, lightweight models for dementia screening using character-level symbolic features. Mekulu et al. introduced CharMark, a novel steady-state Markov modeling approach that identifies linguistic biomarkers from character transitions in spontaneous speech [7]. Separately, symbolic recurrence analysis has been used to reveal temporal linguistic patterns indicative of cognitive disruption [8].

While initially designed for text generation, GPT-2 and its variants have been adapted for dementia classification tasks. These models offer unique advantages in capturing long-range dependencies in language, which can be useful for detecting subtle cognitive changes [9]

Several studies have investigated the use of LLMs for dementia detection. For example, Agvabor et al. [10] trained GPT-3 to predict dementia from spontaneous speech. The LLM achieved an accuracy of 80% in predicting dementia, which was significantly higher than the accuracy of traditional neuropsychological tests. Another study by Koga et al. [11] evaluated the performance of two LLMs, ChatGPT and Google Bard, in generating diagnoses in clinicopathological conferences of neurodegenerative disorders. The results showed that both LLMs were able to generate accurate differential diagnoses with high agreement with the consensus diagnoses of experienced neurologists. Overall, these evidences suggest that LLMs have the potential to be a valuable tool for enhancing early dementia detection. They can be used to analyze large amounts of data from a variety of sources, and they can identify subtle linguistic changes that may be indicative of early dementia.

The introduction of LLaMA (Large Language Model Meta AI) and its successors has opened new avenues for dementia classification research. These models, trained on vast amounts of data, potentially capture a wider range of linguistic patterns [12].

Other types of language models have also been used for dementia detection. For example, pre-trained language models, such as BERT, have also been used to extract features from speech and text that can be used to predict dementia. The pretrained language model BERT was applied to the ADReSS Challenge Dementia Detection Task and achieved state-of-theart results, even though the model was not trained specifically on dementia data [13]. This suggests that BERT can learn to detect dementia from outside data, which is significant because it demonstrates the potential of pre-trained language models for dementia detection.

Furthermore, Adhikari et al. [14] developed a novel dataset of transcripts from AD patients and control subjects in the Nepali language. They then applied a variety of NLP and ML techniques to extract linguistic features from the transcripts. These features were then used to train a machine learning classifier to distinguish between AD patients and control subjects. The authors achieved an accuracy of 94% in classifying AD patients and control subjects using a Naive Bayes (NB) classifier. This result is promising, as it suggests that NLP and ML techniques can be used to develop effective tools for the early detection of AD in low-resource languages. Another investigation by Orimaye et al. [15] showcased the development of a machine learning model to predict probable AD using linguistic deficits, lexical and *n*-gram features extracted from spontaneous speech transcripts using NLP techniques.

Recent advancements in LLMs, such as GPT-4 and LLaMA-2, have further expanded the capabilities of AI in understanding and generating human language. These models, with their increased parameter sizes and sophisticated training methodologies, offer enhanced performance in capturing the intricacies of human speech, making them even more suitable for applications in medical diagnostics. The integration of domain-specific knowledge into these models, coupled with fine-tuning on specialized datasets like DementiaBank’s Pitt Corpus, holds the promise of achieving higher diagnostic accuracy and reliability.

This study builds upon existing research by systematically evaluating and fine-tuning multiple LLMs on a comprehensive dataset, aiming to identify the most effective models for early dementia detection through linguistic analysis. By comparing models of varying complexities and architectures, we seek to understand the strengths and limitations of each, providing insights into their practical applications in clinical settings.

At the core of our research lies the DementiaBank’s Pitt Corpus [16], a comprehensive repository of audio recordings and transcripts from individuals diagnosed with Alzheimer’s Disease (AD) and healthy controls (HC). This corpus provided a robust foundation for our study, encompassing 310 transcripts from 168 AD patients and 242 transcripts from 98 HC subjects. Each participant engaged in the “Cookie Theft” picture description task (see Figure 3), a standard procedure designed to elicit spontaneous speech and uncover underlying cognitive processes [17].

## III. Methodology

This section details our systematic approach to dementia detection using language models. Figure 2 illustrates our end-to-end pipeline, from data preprocessing through model evaluation, designed to ensure robust and reproducible results.

**Fig. 1:**
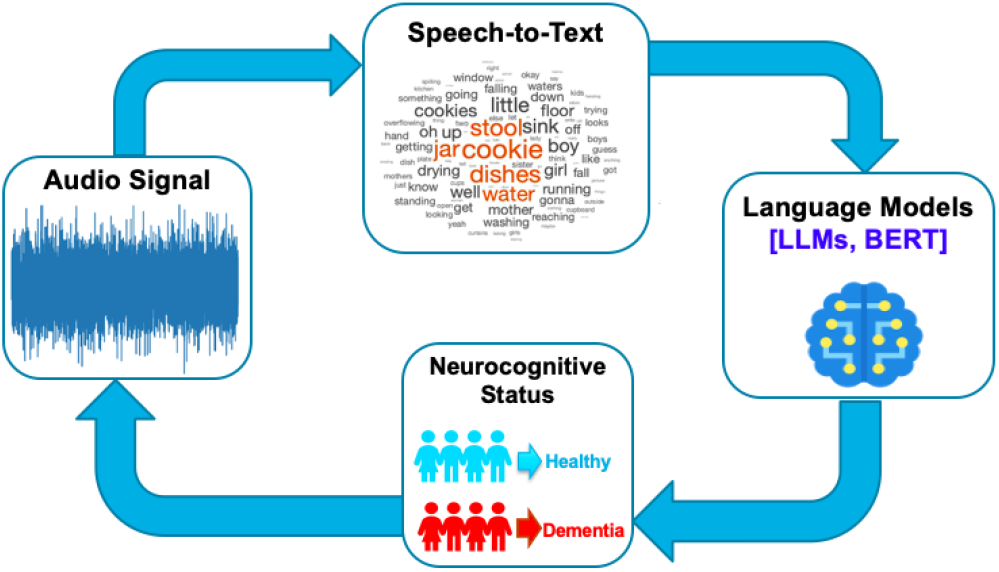
Example of a dementia detection flow diagram using language models.

**Fig. 2:**
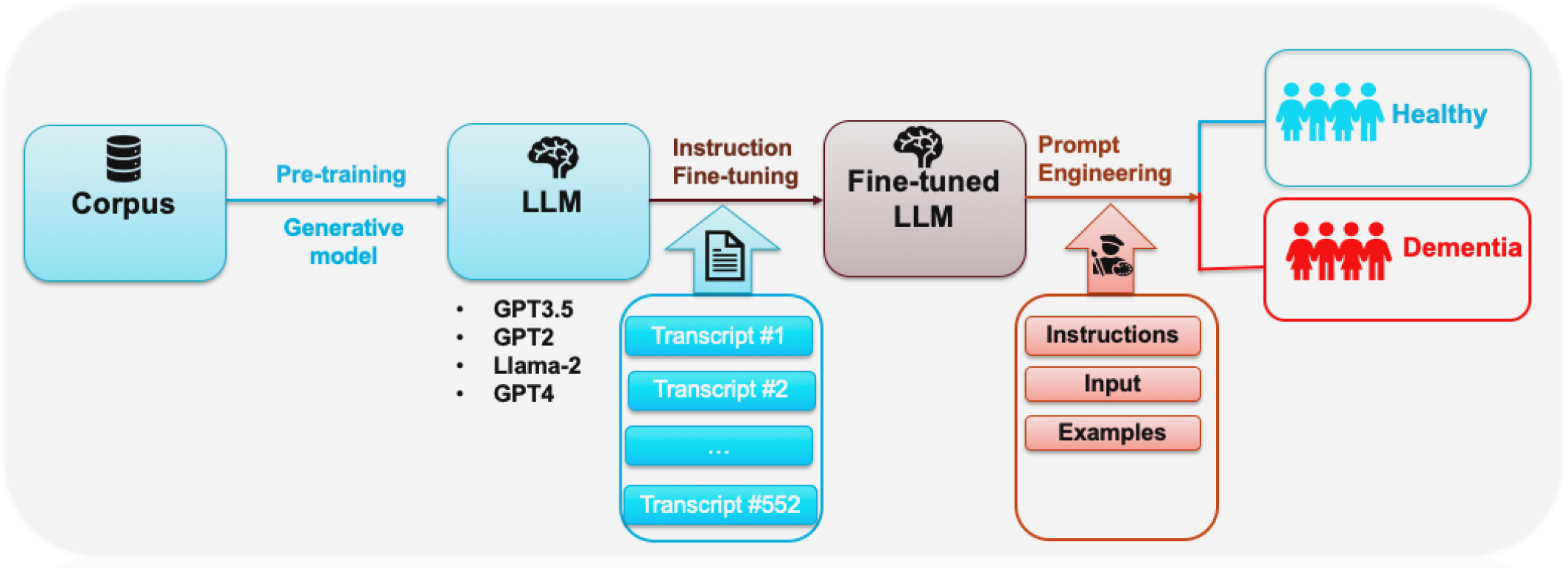
Flow diagram of the proposed methodology.

**Fig. 3:**
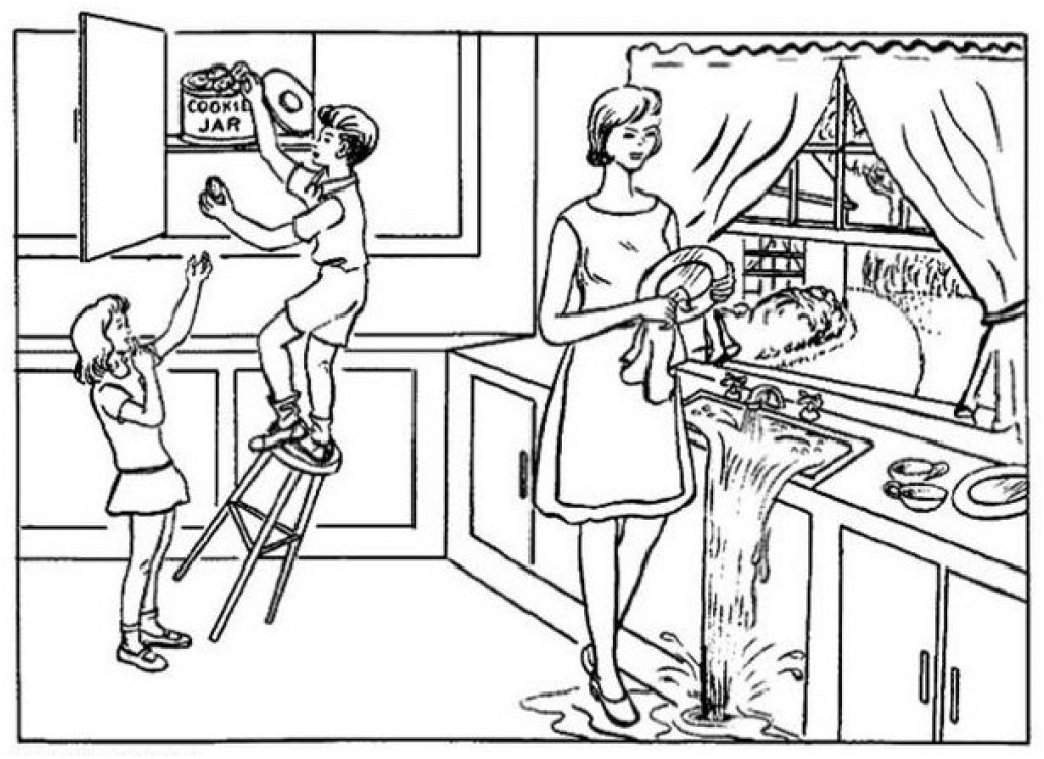
Boston Cookie Theft Picture.

### A. Data Preprocessing

Ensuring the quality and relevance of our data was paramount. We initiated our analysis by preprocessing the transcripts. This involved several key steps:

1. **Transcript Cleaning**: We removed all clinician prompts and questions to focus exclusively on the participants’ natural speech patterns. This purification was essential for capturing authentic linguistic behaviors without external influences.
2. **Text Normalization**: To maintain consistency, all text was converted to lowercase, and punctuation and special characters were stripped away. This standardization minimized discrepancies and facilitated smoother downstream processing.
3. **Retention of Stop Words**: Contrary to common NLP practices, we retained stop words—such as “and”, “the”, and “is”. Preliminary observations indicated that AD subjects frequently exhibit repetitive usage of these words, serving as indicators of cognitive decline [18].
4. **Word Cloud Generation**: To visualize the linguistic landscape, we generated word clouds for AD patients and HC subjects (see Figure 4). These visualizations highlighted the most frequently used words in each group, providing immediate insights into distinct language patterns.

**Fig. 4:**
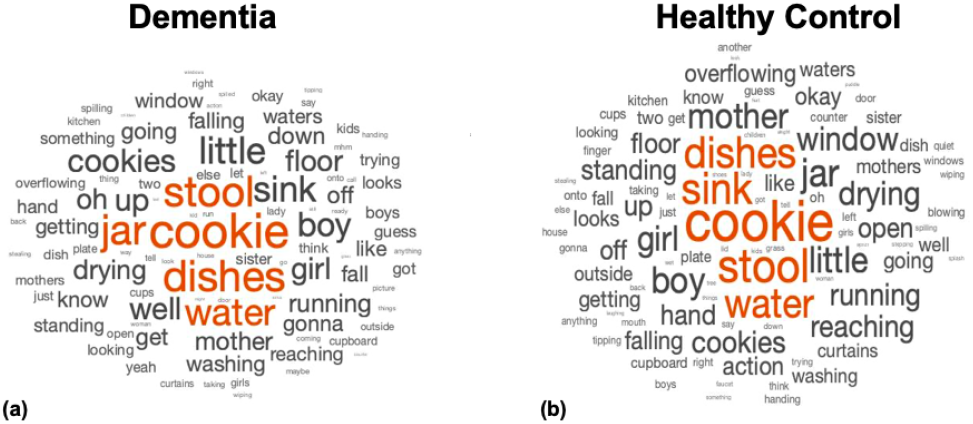
Word clouds for (a) Dementia subjects and (b) Healthy control Subjects.

**Fig. 5:**
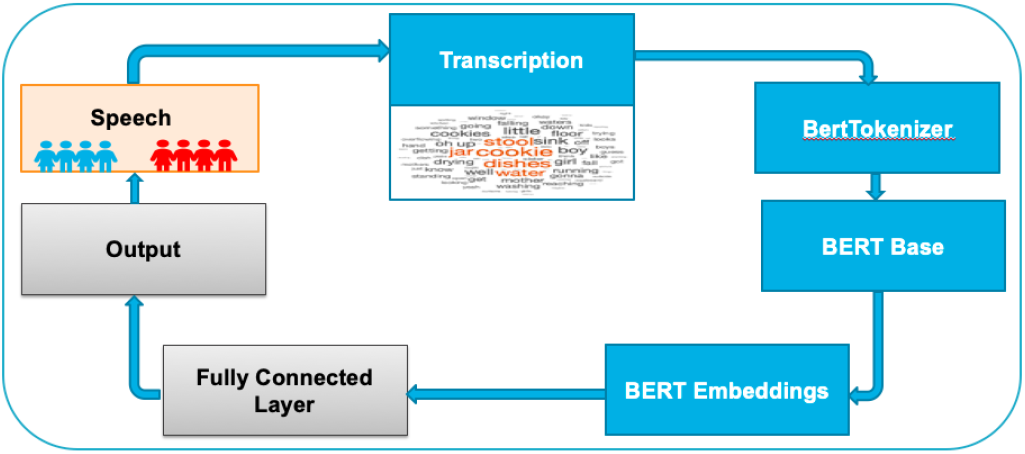
BERT Fine Tuning Flow Diagram.

### B. Computational Models

Our analytical framework leverages both Pre-trained Language Models (PLMs) and Large Language Models (LLMs), each bringing unique strengths to dementia detection.

#### 1) Pre-trained Language Models (PLMs)

**Bidirectional Encoder Representations from Transformers (BERT)** [19] stands out for its deep bidirectional understanding of language context, a crucial capability for detecting subtle linguistic patterns associated with cognitive decline. Unlike unidirectional models that process text either left-to-right or right-to-left, BERT’s bidirectional architecture enables it to capture complex dependencies in both directions simultaneously.

For a given input transcript *X* = [*x*1, *x*2, …, *xn*], BERT first applies WordPiece tokenization and adds special tokens:

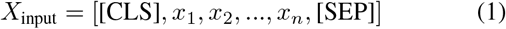

where [CLS] serves as an aggregate sequence representation for classification tasks, and [SEP] marks sequence boundaries. This structured input format enables BERT to learn contextual relationships across the entire transcript.

The model processes this input through L=12 transformer layers (in bert-base-uncased), where each layer combines multi-head self-attention and feed-forward networks:

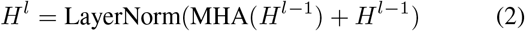

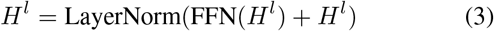

The multi-head attention mechanism, crucial for capturing various aspects of linguistic patterns, is computed as:

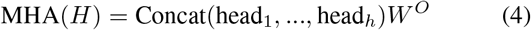

where each attention head focuses on different aspects of the input:

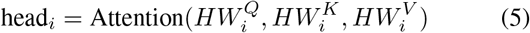

For dementia detection, we leverage BERT’s [CLS] token representation through a classification layer:

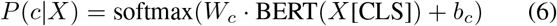

where *Wc* ∈ ℝ*h×*2 and *bc* ∈ ℝ2 are trainable parameters (*h* = 768 for bert-base), and *c* ∈ {0, 1} represents the binary classification outcome. This architecture enables the model to aggregate information across the entire transcript for making diagnostic predictions.

The model’s objective during fine-tuning is optimized using cross-entropy loss:

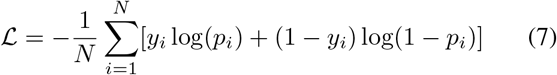

where *N* is the batch size, *yi* is the true label, and *pi* is the predicted probability of dementia. This loss function is particularly suitable for our binary classification task as it penalizes both false positives and false negatives, crucial for clinical applications.

#### 2) Large Language Models (LLMs)

We investigated four state-of-the-art LLMs—GPT-2 (1.5B parameters), GPT-3.5 Turbo (175B parameters) [20], GPT-4 (1.76T parameters) [21], and LLaMA-2 (7B parameters)—for dementia classification. These models leverage transformer-based architectures and extensive pre-training to capture long-range dependencies and subtle semantic patterns in language, characteristics particularly relevant for detecting the linguistic markers of cognitive decline. Due to architectural and accessibility differences, we employed distinct optimization strategies across our LLM implementations. For open-source models (GPT-2 and LLaMA2), we performed complete fine-tuning of model parameters for the dementia detection task. For GPT-3.5 Turbo, accessed through OpenAI’s API, we implemented few-shot learning [22] with carefully engineered prompts. GPT-4, accessed through its web interface, utilized a zero-shot classification approach [23] where each transcript was analyzed based on the model’s inherent understanding of linguistic patterns associated with cognitive decline.

## IV. Experimental Design

Our experimental design systematically evaluates the performance of Pre-trained Language Models (PLMs) and Large Language Models (LLMs) in classifying dementia through speech transcript analysis. This section details our structured approach, encompassing dataset preparation, model training configurations, and comprehensive evaluation metrics.

### A. Dataset Splitting

Our dataset comprised 552 transcripts (310 from AD patients and 242 from healthy controls). To ensure robust model evaluation, we employed a stratified sampling technique that preserved the original class distribution. The data was split into:

- Training set (80%): 442 transcripts (248 AD, 194 HC)
- Validation set (20%): 110 transcripts (62 AD, 48 HC)

This stratified approach maintained the proportion of AD and HC classes across both sets, mitigating potential class imbalance issues and enhancing model generalizability to unseen data. The validation set was held out during training and used exclusively for performance evaluation.

### B. Model Training and Fine-Tuning

Each computational model—ranging from PLMs like BERT to LLMs such as GPT-2, GPT-3.5 Turbo, GPT-4, and LLaMA2—underwent a fine-tuning process optimized to leverage their unique architectures and strengths.

#### 1) Pre-trained Language Models (PLMs)

We implemented BERT fine-tuning using the “bert-base-uncased” variant with AutoModelForSequenceClassification from the Transformers library. For reproducibility, we initialized all random seeds to 42 across PyTorch, NumPy, and Python’s random library components.

Our implementation pipeline consisted of several key stages. First, we developed a preprocessing workflow that preserved linguistic markers by retaining punctuation and special characters, as these could indicate cognitive state. Text tokenization was performed with a maximum sequence length of 512 tokens, using dynamic padding for batch processing efficiency.

The fine-tuning process utilized a custom training configuration optimized for our binary classification task. We monitored model performance through validation after each epoch, implementing model checkpointing to save the best-performing versions based on accuracy metrics. The training process was configured with the hyperparameters shown in Table I1, chosen to balance model convergence with computational efficiency.

**TABLE 1:** BERT Model Fine-Tuning Training Arguments.

| Parameter | Value |
| --- | --- |
| Number of Epochs | 6 |
| Batch Size (Train) | 8 |
| Batch Size (Eval) | 8 |
| Warmup Steps | 500 |
| Weight Decay | 0.01 |
| Logging Steps | 10 |
| Evaluation Strategy | Epoch |
| Save Strategy | Epoch |
| Load Best Model at End | True |
| Metric for Best Model | Accuracy |

To prevent overfitting, we implemented early stopping with model checkpointing, saving the best model state based on validation accuracy. Training progress was logged every 10 steps, allowing for detailed performance analysis and potential hyperparameter adjustments.

#### 2) Large Language Models (LLMs)

We evaluated a diverse set of LLMs, ranging from 1.5B to 175B parameters, employing model-specific optimization strategies based on architecture and accessibility constraints. For GPT-2 and LLaMA-2, we implemented full model fine-tuning, while GPT-3.5 Turbo was accessed through APIs with prompt engineering. Table II details the implementation specifications for each model, highlighting our differentiated approach to model optimization and deployment.

**TABLE 2:** Implementation Details of Language Models.

| Model | Approach | Implementation Details |
| --- | --- | --- |
| GPT-2 | Fine-tuning | • Custom classification head<br>- 3 training epochs<br>- Batch size = 8 |
| LLaMA-2 | Instruction-tuning | • bfloat16 precision<br>- Device-mapped inference<br>- Structured prompts |
| GPT-3.5 | Few-shot (n=5) | • Temperature = 0<br>- 20s timeout<br>- Example-based prompts |
| GPT-4 | Zero-shot | • Web interface<br>- Consistent prompting<br>- Binary classification |

- **GPT-2**: We adapted the base GPT-2 model for sequence classification using GPT2ForSequenceClassification, implementing a custom PyTorch dataset architecture to handle the unique requirements of dementia detection. Our dataset implementation featured dynamic sequence handling with a fixed block size of 384 tokens, utilizing the model’s EOS token for padding operations. This design ensured efficient processing of variable-length inputs while maintaining consistent batch dimensions throughout training. The training pipeline was optimized with a configuration of three epochs and a batch size of 8 for both training and evaluation phases. We implemented a comprehensive monitoring system with validation checks after each epoch and logging at 10-step intervals. Model selection was automated based on accuracy metrics, incorporating early stopping with checkpointing to prevent overfitting and preserve optimal model states. Text preprocessing followed a systematic approach, employing regex-based cleaning with the pattern ‘[â-zAZ0-9]’ to standardize inputs. This included lowercase conversion and careful handling of special characters and punctuation. Each preprocessing step was validated to ensure it preserved essential linguistic features while removing noise that could impact classification performance. The attention mask generation was specifically designed to handle variable-length inputs, ensuring the model focused only on relevant token sequences during inference.
- **LLaMA-2**: Our implementation utilized the “Llama-2-7b-hf” variant, optimized for efficient inference through bfloat16 precision, which significantly reduced memory requirements while maintaining computational accuracy. The model deployment leveraged automatic device mapping to optimize resource utilization across available hardware. The preprocessing pipeline was designed for robust text normalization, incorporating lowercase conversion, whitespace standardization, and systematic handling of special characters. This comprehensive approach ensured consistent input formatting across all samples, critical for reliable model performance. The cleaned text was then processed using a structured dialogue format designed to elicit binary classification decisions. Our prompt engineering approach followed a human-assistant interaction paradigm, with the explicit task description: “The given speech transcript is either from a healthy subject or a diseased subject. Categorize it as one of them.” This format was chosen to leverage LLaMA-2’s training on conversational data. To maintain efficiency in the inference pipeline, we implemented early stopping with a maximum of three new tokens, sufficient for binary classification responses. The entire process was streamlined using HuggingFace’s Dataset library for efficient data management and model interaction.
- **GPT-3.5 Turbo**: We implemented a few-shot learning approach through OpenAI’s API, utilizing a carefully crafted prompt engineering strategy. The model was initialized with a role-based context that framed it as an assistant analyzing conversation transcripts for signs of cognitive impairment. Our prompt structure incorporated five exemple cases for few-shot learning, with explicit instructions to default to “diseased subject” classification when context proved insufficient. To ensure consistency in results, we set the temperature parameter to 0, enabling deterministic outputs. The implementation required robust preprocessing of transcripts, including regex-based cleaning and text wrapping at 100 characters for optimal prompt formatting. To manage API interactions efficiently, we developed a batched processing system that handled 50 samples per batch, with built-in timeout controls (20 seconds per request) and rate-limiting delays (10 seconds between batches) to ensure reliable data collection. For response processing, we implemented a systematic approach to transform model outputs into binary classifications. This included robust error handling for API failures and consistent label mapping for evaluation purposes. The entire pipeline was designed to maximize throughput while maintaining reliability, with particular attention to OpenAI’s API constraints and best practices for large-scale inference tasks.
- **GPT-4**: Our implementation utilized the web interface with a zero-shot classification approach. For each transcript, we prompted the model to perform linguistic analysis based on its pre-trained understanding of dementia-related language patterns. The prompt structure maintained consistency across all samples, requesting binary classification (healthy/diseased) while leveraging GPT-4’s inherent knowledge of cognitive decline markers in speech. This approach differed from traditional fine-tuning or few-shot learning, instead relying on the model’s built-in capabilities to identify linguistic features associated with cognitive impairment. All interactions were conducted with default temperature settings to balance confidence and variability in the model’s responses.

### C. Evaluation Metrics

To comprehensively assess the performance of each model, we employed clinically relevant evaluation metrics that provide a multifaceted view of their diagnostic capabilities. Let TP, TN, FP, and FN denote True Positives, True Negatives, False Positives, and False Negatives respectively. The following metrics were calculated:

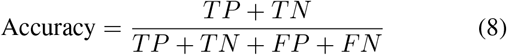

which measures the overall proportion of correct predictions. For clinical performance evaluation:

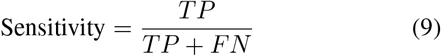

measures the model’s capability to correctly identify patients with dementia (also known as recall or true positive rate), and:

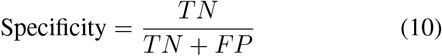

indicates the model’s ability to correctly identify healthy controls (true negative rate). In clinical settings, both sensitivity and specificity *>*= 80% are considered state-of-the-art benchmarks for diagnostic tools [24]. To provide a balanced measure of performance, we calculated the F1-score:

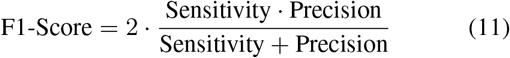

where Precision = TP/(TP + FP) complements sensitivity by measuring the proportion of correct positive predictions.

## V. Experimental Results and Discussion

Our comprehensive evaluation of both PLMs and LLMs revealed distinct patterns in their performance for dementia detection. Table III presents the detailed performance metrics across all models, while Figure 6 provides a visual comparison.

**TABLE 3:** Performance Comparison of Language Models.

| Model | Accuracy (%) | Sensitivity (%) | Specificity (%) | F1-Score (%) |
| --- | --- | --- | --- | --- |
| BERT | 91 | 86 | 95 | 92 |
| LLaMA-2 | 80 | 86 | 89 | 83 |
| GPT-2 | 77 | 82 | 76 | 80 |
| GPT-4 | 73 | 76 | 82 | 78 |
| GPT-3.5 | 50 | 54 | 81 | 65 |

**Fig. 6:**
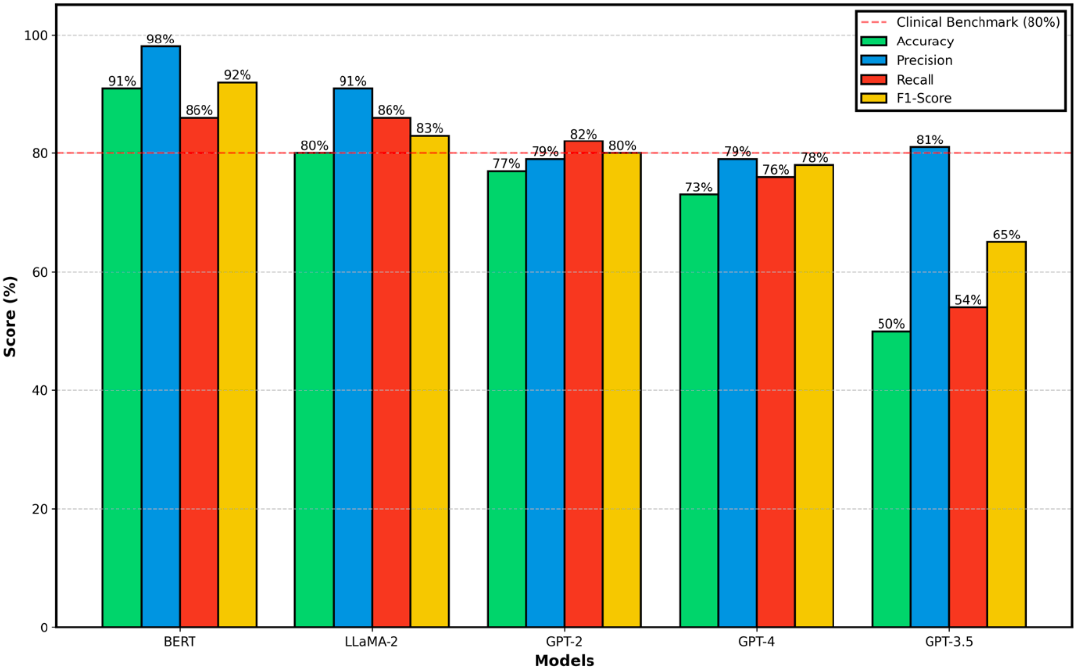
Performance comparison of language models for Dementia detection.

### A. Model Performance Analysis

#### 1) Pre-trained Language Models (PLMs)

BERT demonstrated superior performance across most metrics, achieving the highest accuracy (91%), precision (98%), and F1-score (92%). This exceptional performance can be attributed to:

- Effective capture of bidirectional context in language patterns
- Successful fine-tuning on the specific task of dementia detection
- Robust handling of the structured nature of clinical text classification

#### 2) Large Language Models (LLMs)

Among the LLMs, performance varied significantly:

- **LLaMA-2** emerged as the strongest LLM performer, achieving 80% accuracy and maintaining high precision (91%) and recall (86%). Its strong performance suggests that open-source LLMs can be effectively adapted for clinical applications.
- **GPT-2** showed moderate performance with balanced metrics (accuracy: 77%, F1-score: 80%), demonstrating the capability of smaller LLMs in specialized tasks.
- **GPT-4** achieved respectable performance (accuracy: 73%, F1-score: 78%) despite being used in a prompt-based approach rather than fine-tuning.
- **GPT-3.5 Turbo** showed lower performance (accuracy: 50%), potentially due to limitations in the prompt-based approach for this specific clinical task.

### B. Clinical Relevance and State-of-the-Art Comparison

In the context of clinical dementia detection, sensitivity (recall) and specificity at or above 80% are considered state-of-the-art benchmarks. Our results demonstrate that several models meet or exceed these clinical standards:

- **BERT** achieves 86% sensitivity and 95% specificity, surpassing clinical benchmarks
- **LLaMA-2** matches the clinical standard with 86% sensitivity and 89% specificity
- **GPT-2** shows promising clinical utility with 82% sensitivity, though specificity (76%) falls slightly below the benchmark

These results are particularly significant as they demonstrate that language models can achieve clinically relevant performance levels, potentially supporting earlier and more accurate diagnoses.

### C. Key Findings

Several important insights emerge from our experimental results:

**PLM Superiority in Structured Tasks:** BERT’s outstanding performance validates the effectiveness of PLMs in structured clinical classification tasks, particularly when finetuned on domain-specific data. **LLM Adaptability:** The strong performance of LLaMA-2 demonstrates that larger language models can be effectively adapted for specialized medical tasks when proper fine-tuning is possible. **Prompt Engineering Limitations:** The relatively lower performance of GPT-3.5 Turbo and GPT-4 highlights the challenges of using prompt-based approaches compared to full model fine-tuning in clinical applications. **Clinical Performance Metrics:** Multiple models in our study achieved sensitivity and specificity above 80%, meeting established clinical standards for diagnostic tools. This is particularly noteworthy for BERT and LLaMA-2, which maintained high performance across all clinical metrics. **Precision-Recall Balance:** All models demonstrated varying trade-offs between precision and recall, with BERT achieving the most balanced performance while maintaining clinically relevant thresholds.

These results suggest that while PLMs currently offer superior performance for structured clinical tasks, both PLMs and LLMs can achieve clinically viable performance levels. The findings emphasize the potential for these AI models to complement existing diagnostic tools in clinical settings, particularly given their ability to meet or exceed established clinical performance benchmarks. Furthermore, the strong performance across multiple models suggests robustness in the approach, strengthening the case for their integration into clinical practice.

## VI. Conclusions

This study presents a comprehensive evaluation of both Pretrained Language Models (PLMs) and Large Language Models (LLMs) for automated dementia detection through linguistic analysis. Our findings demonstrate that while both model types show promise, PLMs—specifically BERT—currently offer superior performance for this specialized clinical task. The key conclusions drawn from our research are:

1. BERT achieved exceptional performance with 91% accuracy, 98% precision, and 86% recall, surpassing traditional clinical benchmarks (80%) and establishing a new standard for automated dementia detection. This performance suggests that smaller, task-specific models can outperform larger, more general-purpose language models in specialized clinical applications.
2. Among LLMs, LLaMA-2 emerged as the strongest performer, achieving 80% accuracy and maintaining clinically viable sensitivity and specificity. This finding is particularly significant as it demonstrates the potential of open-source LLMs in healthcare applications when properly fine-tuned.
3. The performance gap between fine-tuned models (BERT, LLaMA-2) and prompt-based approaches (GPT-3.5 Turbo, GPT-4) highlights the importance of task-specific optimization in clinical applications. This suggests that while prompt engineering offers flexibility, fine-tuning remains crucial for achieving optimal performance in specialized medical tasks.

Our research has important implications for the future of automated cognitive assessment. The strong performance of these models, particularly in meeting or exceeding clinical benchmarks, suggests that AI-driven tools could serve as valuable screening mechanisms in clinical settings, enabling earlier detection and intervention in dementia cases. In conclusion, while both PLMs and LLMs demonstrate potential for dementia detection, our findings suggest that carefully fine-tuned PLMs currently offer the most promising path forward for clinical applications. As these technologies continue to evolve, they may increasingly serve as powerful tools to support clinicians in early dementia detection, ultimately contributing to improved patient outcomes through earlier intervention and more accurate diagnosis.

## Data Availability

The data that support the findings of this study are available from the Pitt Corpus within DementiaBank, maintained by TalkBank at Carnegie Mellon University and the University of Pittsburgh School of Medicine. Access to the data in DementiaBank is password protected and restricted to members of the DementiaBank consortium group. In accordance with TalkBank rules, any use of data from this corpus must be accompanied by appropriate corpus references and acknowledgment of grant support (NIA AG03705 and AG05133). Interested researchers can request access to the Pitt Corpus by visiting https://dementia.talkbank.org/access/English/Pitt.html. Established researchers and clinicians working with dementia can contact TalkBank to request access credentials.

## Acknowledgments

The authors of this work would like to acknowledge the NSF grants IIS-2302834 and MCB-1856132 for funding this research.Any opinions, findings, or conclusions found in this paper originate from the authors and do not necessarily reflect the views of the sponsor.

## Notes

### Competing Interest Statement

The authors have declared no competing interest.

### Funding Statement

The NSF funded this study.

### Author Declarations

The study used ONLY openly available human data that were originally located at: https://dementia.talkbank.org/access/English/Pitt.html

